# Creating a lung ultrasound information score to measure the information content of point-of-care lung ultrasound reports

**DOI:** 10.64898/2026.10.30.26366367

**Authors:** Brianna Dinh, Tatiana Vidal, Andrea Hankins, Samuel Lam, Paul Walsh

**Author notes:** **Funding** Pediatric Emergency Medicine Research Foundation Sutter Medical Center Sacramento Foundation.

## Abstract

**Background:** Designing point-of-care lung ultrasound reporting interfaces requires methods for comparison of the information content of the reports generated.

**Objective:** We developed an information score that quantifies the detail in the sonographer’s report. This is distinct from ultrasound severity scores, which reflect disease severity.

**Methods:** Our subject matter experts created a criteria-based, regex-assignable ultrasound report information score. This score was designed to measure the quality of the ultrasound report, with weighting for important clinical findings while retaining its distinctness from ultrasound severity scores. We then tested this score using a publicly available dataset of point-of-care lung ultrasounds performed in a single pediatric emergency department between 30/November/2019 and 14/August/2021. We iteratively refined the initial code we had written with Claude to create a Stata command packaging a set of regex commands that assigned the information score as intended. We compared its results with ultrasound severity and measured its relation with Guiraud’s R, a measure of lexical diversity.

**Results:** We were able to score 415 of the 427 ultrasounds by code; of these, 220/415 (53%) were scored as zero (normal or almost normal). Patients ranged in age from 14 days to 21 years. The score behaved as intended when tested using the dataset. It t retained its distinctiveness from the ultrasound severity score while not being wholly divorced from it. The lung ultrasound information score also correlated modestly with lexical diversity while not being unduly affected by report length.

**Conclusion:** Our information score is an objective, regex-assignable method for comparing information content in point-of-care lung ultrasound reports.

## Introduction

Documenting the findings of point-of-care lung ultrasounds in the pediatric emergency department is generally considered good practice. This ideally comprises a written report and pertinent images. Competing strategies for the design of how the written report component is generated include typed or dictated reports and point-and-click interfaces. Comparing these approaches would be facilitated by developing report information scoring tools for point-of-care lung ultrasound (LUS) reports.

Such a tool is distinct from the many described LUS severity scores^1–5^. Whereas ultrasound severity scores attempt to collapse multiple diverse distinct LUS findings into a single number that predicts clinical outcome, the report information score attempts to collapse the prose of the ultrasound report into a single number that describes the level of descriptive detail in the sonographer’s report. An information scoring tool provides a quality measure of the ultrasound report, ideally with weighting for more severe findings but simultaneously allows good scores for reports that provide good descriptions of less severe findings.

Consequently, an information score inhabits an odd ‘Goldilocks’ statistical space. It should have neither an overly strong nor an overly weak correlation with clinical outcomes, and it should be objectively scored, even though it lacks any kind of gold standard. Whereas ultrasound severity scores are designed for clinicians and researchers, ultrasound information scores are designed for operations (including reporting software design) and process management.

## Methods

This study was approved by our Institutional Review Board. This was a secondary use of a publicly available dataset.^6^ The dataset comprised point-of-care lung ultrasounds performed in a single pediatric emergency department between 30/November/2019 and 14/August/2021. The dataset included 427 lung ultrasounds that had been manually reviewed and assigned an ordinal severity score, along with blood tests, radiology results, and whether the patient was admitted or discharged. In this dataset, the lung ultrasounds were performed prior to other ancillary testing; laboratory testing and imaging were performed based on the clinical judgment of the physician rather than mandatory protocols, and ultrasound severity scoring had been performed based on the ultrasound report alone (blinded to other data).

In the first part of this work, our subject matter experts initially created and then refined an information weighting schema. This schema was designed to reflect how much the sonographer described, weighted by more worrisome findings that might provoke additional investigations or hospitalization.

The scoring scheme is summarized in **Table 1** and described in detail in the supplemental materials. Certain findings in the narrative body of the ultrasound report on their own may trigger additional testing or more aggressive management: we assigned these a score of 15; findings that may support hospitalizing the child were assigned a score of 20. We also added points for descriptions suggesting increased severity; for example, pneumonia received additional weighting when it was multilobar, bilateral, or accompanied by a pleural effusion.

**Table 1.** Information score and descriptions of abnormalities.

| Domain | Findings and wording counted | Points |
|---|---|---|
| **Vertical artifacts (B-lines) [maximum attained]** |  |  |
| B-lines | None | 0 |
|  | Borderline: few, occasional, isolated, scant, rare, sparse, minimal, trace, scattered, several, "a small number of", "increased" B-lines in one area | 1 |
|  | Patchy or multifocal B-lines; multiple, several, scattered or numerous areas; excessive short B-lines | 3 |
|  | Carpet-like B-lines | 5 |
|  | Excess, long or diffuse B-lines; B-lines in all zones | 10 |
|  | Coalescent or confluent B-lines; white-out | 15 |
|  | A limiting word (occasional, few, scant, rare, sparse, isolated) before excess/diffuse/numerous caps the score at 1. | note |
| **Areas: number of sub-zones with B-lines [added only when B-lines score 5 or more]** |  |  |
| Sub-zones named | Upper, mid or lower posterior; posterior; base; axillary; anterior; diaphragm. "All zones" counts as five. Each counted once. Windows scanned ("interrogated") do not count, only where findings are described. | 1 zone: +0<br>2: +5<br>3: +7<br>4: +10<br>5: +15 |
| **Pleura [maximum attained: pleural pathology term x quantifier]** |  |  |
| Pleural pathology | Moth-eaten, irregular, string of pearls, punched-out, thickened or edematous pleura | 2 if no quantifier |
| Pleural quantifier | Very mild, minimal, trace, tiny, slight | 1 |
|  | Mild, or none stated | 2 |
|  | Mild to moderate | 4 |
|  | Moderate | 7 |
|  | Moderate to severe | 15 |
|  | Severe | 20 |
| **Air bronchograms and parenchyma [maximum attained, except air bronchograms add to pneumonia]** |  |  |
| Air bronchograms | Air bronchograms, not otherwise specified (including linear) | 5 |
| Atelectasis | Atelectasis | 5 |
| Pneumonitis, bronchiolitis | Scored alike, by quantifier: very mild 2; mild or none stated 5; mild to moderate 7; moderate 10; moderate to severe 12; severe 15 | 2 to 15 |
| Pneumonia pattern | Pneumonia, consolidation, shred sign; air bronchograms described as concentric, irregular, static or clustered; C-stripe, C-line, cupola, dome | 15 |
|  | Extensive pneumonia (multilobar, bilateral, both lungs, extensive, severe); cauliflower pattern; hepatization | 20 |
| Air bronchograms with pneumonia | When pneumonia or consolidation also scores, air bronchograms add to it: mentioned / described as concentric, irregular, static or clustered | +5 / +10 |
| **Pleural effusion [scored per side]** |  |  |
| Size | Tiny, trace or minimal 5; small 10; moderate 15; large or extensive 20; no size stated 10 | 5 to 20 per side |
| Sides | Left and right both named: sum of the two sides. "Bilateral" alone: twice the size score. One side: that side only. | sum / x2 |
| Loculation | Loculated or non-loculated | +5 |
| With pneumonia | The larger of the above and the pneumonia tiers: tiny 5; small 15; moderate or large 20 | 5 / 15 / 20 |
| **Side and/or location [only when an abnormal finding scores; both may apply]** |  |  |
| Laterality | Left, right, bilateral, unilateral, bibasal; or "all zones" | 3 |
| Sub-zone localization | Any named sub-zone (as above) in a finding clause; or "all zones" | 2 |
| **Etiology and differential [maximum 15 in total]** |  |  |
| Etiology or differential | Any specified etiology (for example COVID-19, RSV, influenza, adenovirus, bacterial, aspiration, cardiogenic edema), or a differential diagnosis supplied (differential, versus, consider). The two do not add. | 15 |
| Not counted | "Viral" alone; asthma or reactive airway; "bronchiolitis versus pneumonitis"; indication field; text after the signature | 0 |
| **Cardiac windows** |  |  |
| Cardiac | Any cardiac finding, including normal | 1 |
| **Other** |  |  |
| Pneumothorax | Not scored in the default analysis (optional points- recommended default 20 for medical cases, higher for Trauma. | 0 (20) |
| Normal impression | Impression normal, unremarkable, within normal limits, or negative: total score | 0 |
| Missing | Blank report, or an unfilled template | missing |

The information score is the sum of the components above. Where a component says maximum attained, only the highest matching tier is counted. Findings that are negated in the report (for example, "no effusion") are not scored.

Where the ultrasound was normal, the information score was set to zero. This is because no amount of narrative eloquence adds clinical value to a normal imaging study. In a survey of clinicians, the single word ‘normal’ was considered an adequate report for a normal screening chest X-ray.^7^ Non-patient-specific text describing the machine, probe, and image storage details was not counted towards information scoring.

The score was designed to be assignable by regex code, a set of computer commands to parse text and extract phrases, to avoid reviewer bias and facilitate automation in larger datasets. One author (PW) developed and refined the information scoring tool using an iterative process with SL, an author with pediatric emergency medicine and ultrasound expertise.

We initially attempted to create the regex code using a large language model (LLM), Perplexity, but this was unsuccessful, so the initial regex code was almost all handwritten. After initially demonstrating the feasibility of doing this study using only the handwritten code, we again attempted to use another LLM (Claude, Anthropic) to refine our code. We manually checked the performance of the LLM model against the handwritten code on 200 lung ultrasound reports. We iteratively corrected the LLM code and re-tested it until it was producing subjectively better results than the handwritten code. We then used the LLM to package this code into a Stata command that others can easily use. Cases whose ultrasounds could not be assigned a score were compared with those that could.

In the second part of this work, we validated the observed versus the expected behaviour of this information scoring scheme against an available dataset of pediatric point-of-care lung ultrasounds. ^6^

There were three components to the validation. First, the information score would be expected to have a weak but non-zero correlation with the severity of the ultrasound score previously assigned. Second, the information score would be expected to correlate with three groups defined by its thresholds, describing discharge without laboratory testing; discharge following laboratory testing; and those who were hospitalized. However, this correlation should be diminished or disappear when adjusted for severity of the ultrasound findings. Third, we would expect a modest but non-zero correlation between the information score and the lexical richness of the ultrasound report.

We correlated the information scores with the severity of the ultrasound score previously assigned using Spearman’s rank correlation on abnormal ultrasounds. We attempted ordinal logistic regression of the information score against ultrasound severity category and tested the underlying assumptions of our ordinal regression models using the Brant and

Wolfe-Gould tests and Wald tests.^8–10^ We performed quantile (and ordinary least squares) regression and estimated the proportion of the information score not attributable to ultrasound severity as 1-(pseudo)R^2^.

We compared the information scores to the three groups defined by the thresholds describing discharge without laboratory testing and chest X-rays, discharge following laboratory testing and chest X-rays, and those who were hospitalized. We used generalized ordered logistic regression to model the information score against these three outcomes with and without adjustment for ultrasound severity. We anticipated a modest relationship in the unadjusted regression that would disappear following adjustment for severity. The decision to obtain blood tests or hospitalize rests on many patient, disease, and social characteristics in addition to the lung ultrasound. We would not expect an information score measuring a lung ultrasound’s information content to serve as a clinical prediction rule. Therefore, we omitted some of the modelling commonly performed for clinical prediction rules.

We then calculated Guiraud’s R for each ultrasound report and compared it with the lung ultrasound information score using Spearman’s *rho*. ^11^ We plotted boxplots of Guiraud’s R and length for each grade of ultrasound severity. is used in applied linguistics as a measure of lexical diversity, a component of lexical richness, that corrects for text length. It is calculated by dividing the total number of unique words by the square root of the total word count to prevent text length from being mistaken for lexical richness. Higher scores imply greater lexical diversity (a native student essay might have a Guiraud’s R of 6.5-8.0, professional literature 10.0-20.0+, a draft of this manuscript had a Guiraud’s R of 14.3).

## Results

We successfully created and packaged as a Stata command a set of regex commands that assigned the information score as intended. We were able to score 415 of the 427 ultrasounds by code; of these, 220/415 (53%) were scored as zero (normal or almost normal). Patients ranged in age from 14 days to 21 years. The severity categorization and prevalence of abnormalities are shown in **Table 2**. Among abnormally reported scans, the mean information score was 22 (SD 13), and the median was 20 (IQR 13, 32). These are shown in **Figure 1**.

**Table 2.**
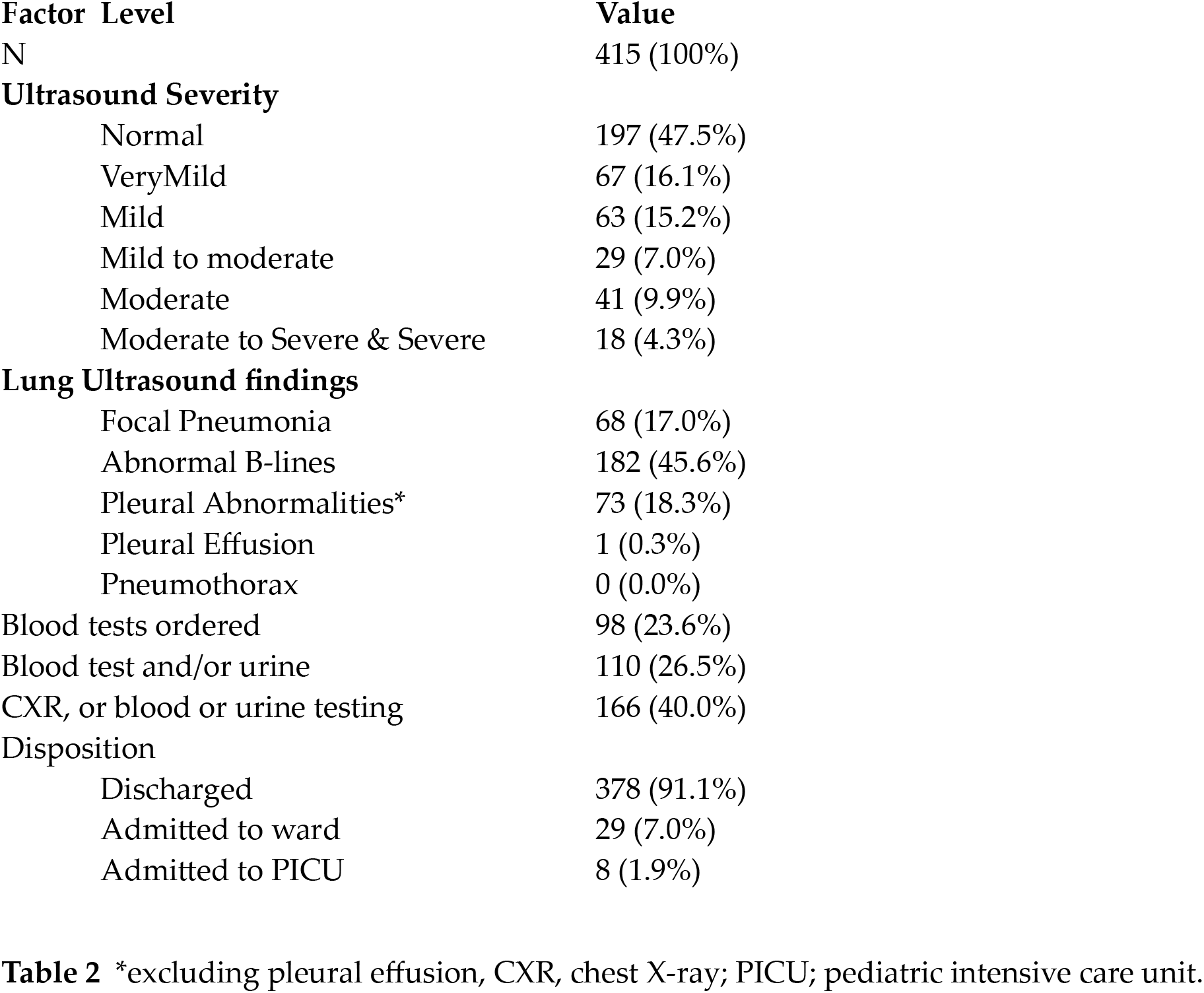
Sample characteristics.

| Factor | Level | Value |
|---|---|---|
| N |  | 415 (100%) |
| **Ultrasound Severity** |  |  |
|  | Normal | 197 (47.5%) |
|  | VeryMild | 67 (16.1%) |
|  | Mild | 63 (15.2%) |
|  | Mild to moderate | 29 (7.0%) |
|  | Moderate | 41 (9.9%) |
|  | Moderate to Severe & Severe | 18 (4.3%) |
| **Lung Ultrasound findings** |  |  |
|  | Focal Pneumonia | 68 (17.0%) |
|  | Abnormal B-lines | 182 (45.6%) |
|  | Pleural Abnormalities* | 73 (18.3%) |
|  | Pleural Effusion | 1 (0.3%) |
|  | Pneumothorax | 0 (0.0%) |
| Blood tests ordered |  | 98 (23.6%) |
| Blood test and/or urine |  | 110 (26.5%) |
| CXR, or blood or urine testing |  | 166 (40.0%) |
| Disposition |  |  |
|  | Discharged | 378 (91.1%) |
|  | Admitted to ward | 29 (7.0%) |
|  | Admitted to PICU | 8 (1.9%) |

\*excluding pleural effusion, CXR, chest X-ray; PICU; pediatric intensive care unit.

**Figure 1.**
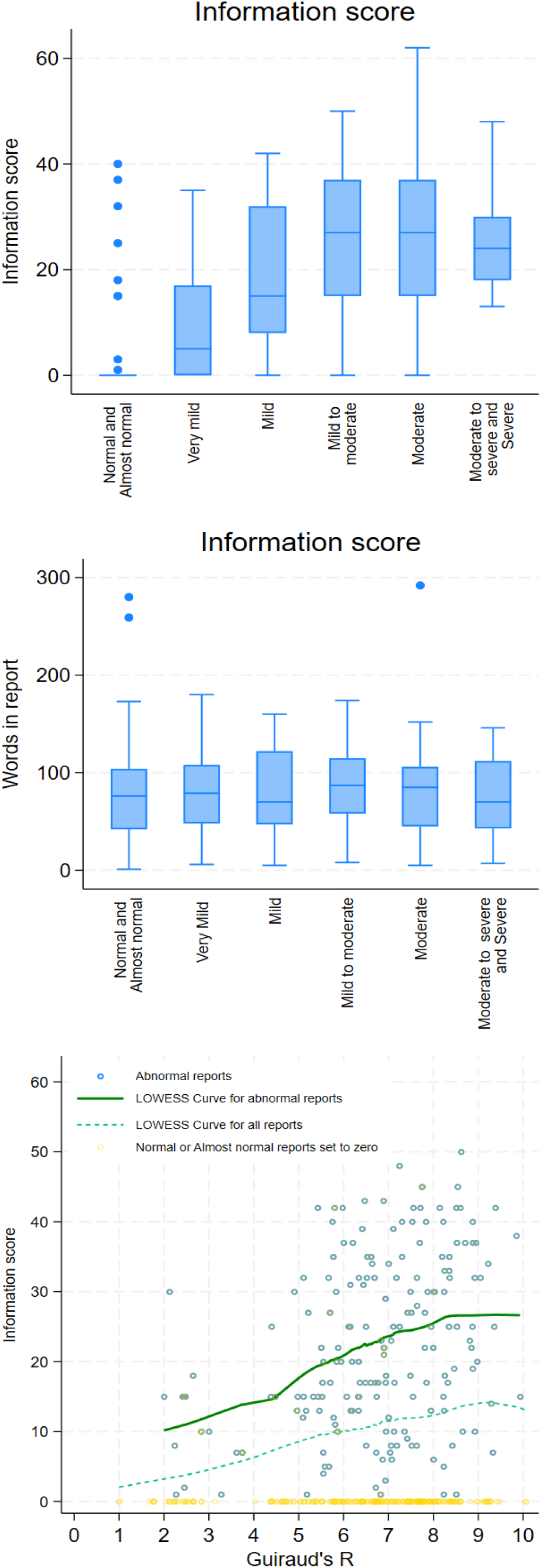
The top panel shows the wide range of information scores across ultrasound severity classes, whereas report length does not vary much between severity classes. The bottom panel shows that lexical diversity increases with score, then plateaus.

The proportional odds assumption inherent ot ordinal logistic regression was not met for models regressing the ultrasound severity score and lung information score, although the individual ORs at each severity level were significant. The unadjusted pairwise correlation between the total information score and the manual severity categorization was 0.70, *p* < 0.0001. Quantile regression (ultrasound severity score was heavily skewed towards normal) showed a coefficient of 0.94 (95% CI 0.088, 0.099; *p* <0.001) with pseudo R^2^ 0.43 (R^2^0.49 in OLS), leaving 57% of the information score not being explained by ultrasound severity, indicating that the scores are related but distinct. This is to be expected because we set normal and almost normal ultrasounds to have an information score of zero. Excluding normal studies, the pseudo R^2^ was 0.124, further emphasizing that the information and severity scores are distinct.

Ordinal logistic regression showed an odds ratio (OR) of 1.53 (95% CI 1.33, 1.76) for increasing severity of outcome (discharged home without blood tests or a chest X-ray, discharged home following blood tests or a chest X-ray, or admission) for each 10-point increase in the information score. The proportional odds assumption was met (Brant test *p*=0.391).

When adjusted for ultrasound severity, this effect disappeared, OR 1.11 (95% CI 0.91, 1.36; *p*=0.29). This helps demonstrate that ED clinical outcomes and descriptive detail of the report captured in the information score are distinct. Since ED point-of-care lung ultrasound is performed by the treating physician, this is unsurprising, but this analysis does not address outcomes caused by communication gaps with admitting services that could arise if the ultrasound report was inadequate.

We observed a wide spread of information scores across categories despite relatively similar report length. We found a modest correlation (Spearman’s rho 0.125, p=0.011 ) between the lexical richness of the ultrasound reports, measured using Guiraud’s index, and the lung information score. The raw data with an overlaid LOWESS curve is shown in **Figure 1**.

Patients whose ultrasound results (N=12) could not be coded using regex code were more likely to have pneumonia (*p*=0.007) and abnormal B lines *(p*=0.04) on ultrasound and to be

## Discussion

Our point-of-care lung ultrasound information score behaved as it should in this validation dataset. Because the lung information score is weighted, it should demonstrate some correlation between severity and threshold clinical outcomes, discharging without additional testing, discharging after additional testing, and hospital admission. The information score should nonetheless remain distinct from ultrasound severity scores. Because the information score is measuring text, it should increase with increasing lexical diversity up to a point where additional unique words do not add new information.

The information score differs from clinical scoring systems as the aim is not to predict clinical outcomes, but rather to measure the quality of radiology reporting. These are not unrelated, and so some correlation between clinical outcomes and the information score would be expected, and this is what we observed. However, information scores have the potential to be high where multiple findings in multiple locations are described in detail, but none of them alone or in combination is enough to cause severe clinical illness. Conversely, a report can be very pithy and concise in severely ill patients. We observed both of these phenomena in our data.

The role of information scores such as this one is in the realm of systems design, including software used to report point-of-care lung ultrasounds, and operations management rather than clinical care. Such tools live in the somewhat self-contradictory space of needing to value quality reporting of severe outcomes more than non-severe ones while not penalizing good quality reporting of less abnormal ultrasounds; and they represent a compromise of sorts. (Physician readers will recognize this from mandates to simultaneously spend more time with each patient and increase the number of patients they see per hour.)

## Limitations

The public use dataset covered a period of higher SARS-CoV-2 prevalence than subsequent periods when this scoring system may be applied. This marked the beginning of a period of increased point-of-care lung ultrasound in this department, and reporting has subsequently gradually improved with greater use of clearly identified and often structured template-based reports. We addressed this by retaining the originally defined thresholds and avoiding overfitting by optimizing anchor thresholds based on the validation dataset. This dataset also (correctly) assumes imaging and blood tests were ordered because they were thought to be clinically indicated rather than being protocol-driven.

We could not extract and assign an information score to 12 ultrasound reports by code. This reflected limitations of the snippets of charting text from which the reports were extracted in the original paper. These cases tended to be sicker than those patients included in this study, although it is not clear how this would change our results. This also reflects our decision to include only reports that could be assigned a score using regex functions. Manual information score assignment would limit reproducibility and would severely limit the utility of the information score. We speculate that this decision protected our analysis from latent subconscious biases on the part of the authors. Large language models (LLMs) might be an alternative but raise potential difficulties with reproducibility both between different LLMs and even within individual LLMs. Our initial attempts at using LLMs had to be abandoned because of cost.

We did not calculate inter-rater characteristics of our results because the use of regex code to assign information scores meant there could be no differences between repeated runs.

Guiraud’s index has been criticized both as being an overly severe correction for longer texts and as overly simplified compared to alternative measures. However, because ultrasound reports should be inherently brief with limited scope for lexical diversity given their technical nature compared with novels and more usual linguistic materials, we felt that Guiraud’s index was adequate.

The scope of what is being measured should not be overstated. The written lung ultrasound is only one piece of performing point-of-care lung ultrasound. Just as immaculate charts can mask perfunctory care, reviewing point-of-care lung ultrasound reports alone does not substitute for assessing the quality and interpretation of scanning by reviewing images and bedside observation. Similarly, analysts could choose to modify threshold values to local needs.

## Conclusion

The lung ultrasound information score presented here demonstrated appropriate behavior in this validation dataset. It retained its distinctiveness from the ultrasound severity score while not being wholly divorced from it. The lung ultrasound information score also correlated modestly with lexical diversity while not being unduly affected by report length.

## Data Availability

This is a secondary use of a publicly available database from a published article. The data, as reprocessed by us, is available upon request.

## Acknowledgements

The authors gratefully acknowledge the assistance of Heejung Bang PhD for her statistical support.

